# Long-term survival, psychiatric, physiological, and functional outcomes of critically ill patients requiring prolonged mechanical ventilation: a systematic review

**DOI:** 10.1101/2023.11.27.23299077

**Authors:** Jarryd Ludski, Conor Honeywill

**Author notes:** **Corresponding Author:** Dr Jarryd Ludski, School of Medicine Sydney, The University of Notre Dame Australia, 160 Oxford Street, Darlinghurst, New South Wales, 2010, Australia. **Secondary Contact:** Dr Conor Honeywill.

## Abstract

**Purpose:** Invasive mechanical ventilation is utilized in over 50% of Australian Intensive Care Unit patients, with rates increasing world-wide. Prolonged mechanical ventilation is associated with increased length of hospital stay and in-hospital mortality compared with patients ventilated under the time threshold. Previous studies have highlighted mortality rates, but much remains unknown regarding the long-term physiological, functional, and psychiatric effects of prolonged mechanical ventilation. With a greater understanding of these outcomes, models of care can be formulated to reduce long-term morbidity.

**Methods:** Medline, CINAHL and the Cochrane Library were searched between 1998 and March 2019, for PMV, patients in ICU and long-term outcomes. Included studies had patients, that received greater than or equal to 14 days of IMV. Primary outcome was mortality rates with secondary outcomes clustered into physiological, functional, and psychiatric outcomes.

**Results:** 1057 studies were identified, with 24 included. 73% of PMV patients were discharged from ICU, with mortality rates of 57% and 69% at 12 and 48 months. 30.2% were discharged home, 25% developed new onset ventilator acquired pneumonia and up to 39% experienced psychiatric complications.

**Conclusion:** Despite a high proportion of patients surviving to hospital discharge, subsequent outcomes are extremely poor for patients that require PMV.

## BACKGROUND

Invasive mechanical ventilation (IMV) is used to generate a controlled flow of gas into a patient’s airways, aiming to improve the ventilation-perfusion (V/Q) matching and reduce respiratory effort. It is recognized as one of the most common interventions in intensive care units (ICU), with a point prevalence of 58% of patients in Australian and New Zealand ICUs (Rose and Gerdtz 2009). Globally, epidemiological trends of the use of IMV have increased significantly since 1993 (Mehta et al. 2015), along with the incidence of prolonged mechanical ventilation (PMV). A study based in the United Kingdom reports IMV in 4.4 cases per 100 ICU admissions, with 6.3 cases per 100 ventilated ICU patients meeting criteria for PMV (Lone and Walsh 2011). Whilst the definition of PMV varies within the literature, a review looking at 419 studies identified greater than 14 days and 21 days as the most consistent definitions (Rose et al. 2017).

PMV is important to define, as those who receive the intervention have a significantly increased length of hospital stay and in-hospital mortality when compared to those who are mechanically ventilated under the time threshold (Lone and Walsh 2011). A meta-analysis which investigated prolonged mechanical ventilation, defined as greater than 14 days, found a pooled in-hospital mortality rate of 29%, with a dire one-year mortality rate of 59% (Damuth et al. 2015). Additionally, they demonstrated significant morbidity to the patient and burden on the healthcare system, with only 19% discharged home. Moreover, PMV is associated with high resource utilization, and greater costs to the healthcare system when compared to non-prolonged mechanically ventilated patients (Lone and Walsh 2011; Cox et al 2007).

Although long-term mortality has been investigated, little is known of the long-term physiological, functional, and psychiatric effects of PMV. It is described in the literature, that patients experiencing PMV are at risk of psychiatric morbidity and a decreased quality of life (Oeyen et al. 2010; Hung et al. 2010). Studies have also shown symptoms suggestive of post-traumatic stress disorder, anxiety and mood disorders following the intervention (Rose et al 2014).

Physiologically, PMV patients may be predisposed to developing, respiratory muscle weakness, low tidal volumes and a high respiratory rate (Purro et al. 2000; De Jonghe et al. 2007). Whilst the associated prolonged bed rest may result in peripheral muscle weakness or critical illness polyneuropathy (Zhou et al. 2014), leading to poorer physiological and functional outcomes (De Jonghe et al. 2007; Chen et al. 2012).

Questions regarding how these symptoms and effects change over time, whether resolving or worsening, remain unanswered. This population is already recognized to be at an increased risk of morbidity and mortality, however a greater understanding of the long-term outcomes is essential for the true impact of PMV to be assessed and mitigated.

### Objectives of the study

We aim to highlight the prevalence and course of these outcomes following ICU discharge and into the future including survival rates and rates of physiological, functional, and psychiatric outcomes. Furthermore, we hope this understanding will help to guide appropriate follow-up care strategies to ameliorate these burdens. Post-ICU follow-up models have already demonstrated improvements in psychiatric and functional outcomes in patients **^14^** however limited findings have been produced in the PMV population.

## METHODS

### Search strategy and selection criteria

Study protocol was written in accordance with Cochrane handbook guidelines and the Preferred Reporting Items for Systematic Reviews (PRISMA) statement. We searched MEDLINE, CINAHL and Cochrane databases for publications in the last 20 years, from 1998 to March 10 2019, with the following search terms and their related terms: “Intensive care unit”, “Prolonged mechanical ventilation” and “long-term outcome”. **Appendix A** displays the search design. No original language restrictions were set. Additional papers were added following reference tracking and search results were merged using EndNote with duplicates removed. Studies were included if they were available in English and reported outcomes at greater than 3-months post discharge of patients over 18 years, admitted to an intensive care unit (ICU), coronary care unit (CCU) or equivalent who received 14 days or greater of mechanical ventilation. If a studies original design did not explicitly meet our inclusion criteria, but their final reported population and outcomes did, the study was included. We excluded studies where mechanical ventilation was resulting from solid organ transplantation or traumatic brain injury, given likely confounding of morbidity and mortality data. Correspondence, and editorials were excluded.

### Data assessment and abstraction

The initial relevance screen of titles and abstracts was performed by two reviewers independently (JL, CH), with results categorised into irrelevant results and relevant results or not possible to determine. Exclusion logs were compared, and cases of disagreement were included. All studies categorised as relevant or not possible to determine underwent full manuscript review for eligibility within the study criteria by two reviewers independently (JL, CH), with any disagreement discussed with a third contributor (SR).

### Data extraction

Using a standardised data collection form, two reviewers (JL, CH) extracted data for study design, population descriptors, length of mechanical ventilation, survival rates and physiological, function and psychiatric outcomes at all timepoints.

### Outcomes

The primary outcome was post-discharge mortality at 3, 6, 12 and 24 months. Secondary outcomes were clustered to “Physiological”, “Functional” or “Psychiatric” outcomes reported at greater than 3 months post-hospital discharge. Physiological outcomes included development of sepsis, multi-system organ failure, Acute Respiratory Distress Syndrome (ARDS) and Ventilator acquired pneumonia (VAP). Functional outcomes included critical illness polyneuropathy and myopathy (CIPM), Quality of life (QoL) to perform activities of daily living (ADLs), fragility index, exercise tolerance and pulmonary function tests. Psychiatric outcomes included Post-intensive care syndrome, Post-traumatic stress disorder (PTSD), delirium and cognitive impairments. Data was collaborated using pooled analysis.

## RESULTS

1057 studies were identified with 85 passing a relevance screen and undergoing full manuscript review (**see** **figure 1**). 24 papers were included after meeting all inclusion and exclusion criteria. **Table 1** lists the reasons for exclusion. Of the 24 studies, 1 was a systematic review and 23 were cohort studies with 11 prospective and 12 retrospectives. Patient characteristics from the 24 included studies are highlighted in **Tables 2a and 2b**. The study population included 335,703 patients. Median reported age was 64 years, with a mean of 63.98 years (from 16 studies). 45.4% of patients were female.

**Figure 1.**
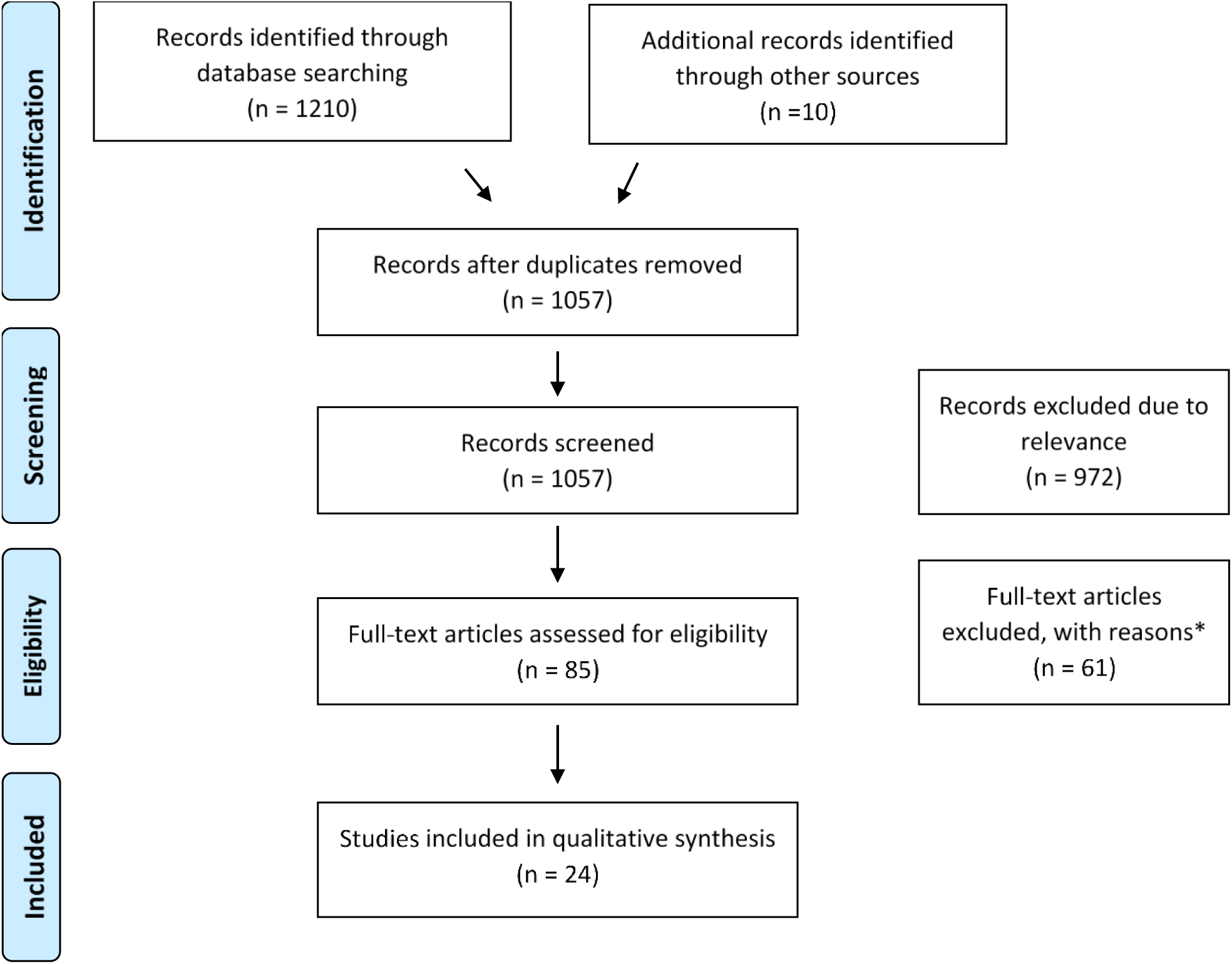
Study flow chart * reason for exclusion outlined in Table 1

**Table 1.** highlights reasons for exclusion.

| Reason for exclusion | Number of studies |
| --- | --- |
| Excluded via relevance screen | 972 |
| Excluded after full manuscript review | 61 |
| Did not meet criteria for prolonged mechanical ventilation | 41 |
| Outcomes not reported at greater than three months | 3 |
| MV result from organ transplant, traumatic brain injury or other excluded causes | 3 |
| A review, commentary, or narrative article | 9 |
| Review not published in English | 2 |
| Unable to access transcript | 2 |
| Patient population not appropriate due to age | 1 |

**Table 2a.**
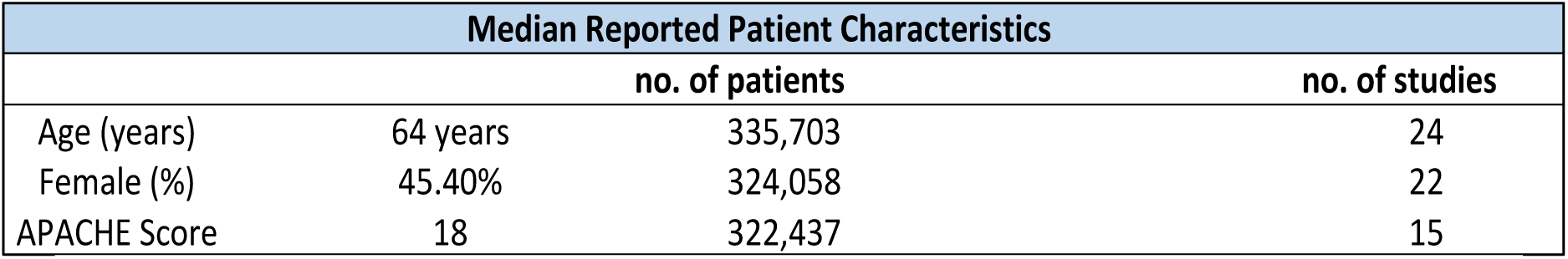
Data are median reported statistics. APACHE = Acute Physiology and Chronic Health Evaluation. Total study population =335,703 patients from 24 included studies.

**Table 2b.** Data are reported means. APACHE = Acute Physiology and Chronic Health Evaluation. Total study population = 335,703 patients from 24 included studies.

| Pooled Mean Patient Characteristics |  |  |  |
| --- | --- | --- | --- |
|  |  | no. of patients | no. of studies |
| Age (years) | 63.98 years | 321,287 | 16 |
| Female (%) | 45.40% | 324,058 | 22 |
| APACHE Score | 19.14 | 320,013 | 8 |

Median Acute Physiology and Chronic Health Evaluation (APACHE) score was reported in 15 publications, calculated as 18, while a mean of 19.14 was calculated from 8 publications. 22 studies reported mortality outcomes, with an in-hospital mortality rate of 27% (from 17 studies, 334,524 patients) and rates of successful weaning from PMV equal to 57% of patients as reported in 10 studies.

Mortality outcomes were extracted, and pooled analysis used to account for varied study populations, with post-discharge mortality rates and total mortality rates reported at 3, 6, 12, 24, 36, 48, 60- and 120-months (outlined in **Table 3a and 3b**). Post-discharge mortality rates of 32% were reported at 12 months up to 70% at 60 months. Total mortality was more commonly reported than post-discharge mortality, with the 12-month timepoint being reported in 20 papers, including 335,456 patients. Total mortality was reported as 57% at 12 months, and 69% at the 48-months. Smaller study populations were reported at 3, 6, 24, 36, 60- and 120-month timepoints.

**Table 3a.** shows pooled reported Total mortality rates and post-discharge mortality rates at intervals since discharge from ICU or equivalent. No studies reported post-discharge mortality rates at 24,36,48 or 120 months.

| Months since discharge | Pooled Total Mortality Rate | n (patients) | Pooled Post-discharge Mortality Rate | n (patients) |
| --- | --- | --- | --- | --- |
| Never discharged | 27% | 334,524 |  |  |
| 3 | 33% | 336 | 21% | 283 |
| 6 | 13% | 12,282 | 31% | 194 |
| 12 | 57% | 335,456 | 32% | 2,341 |
| 24 | 25% | 11,594 | - | - |
| 36 | 49% | 842 | - | - |
| 48 | 69% | 318,621 | - | - |
| 60 | 43% | 12,109 | 70% | 10 |
| 120 | 58% | 11,631 | - | - |

**Table 3b.**
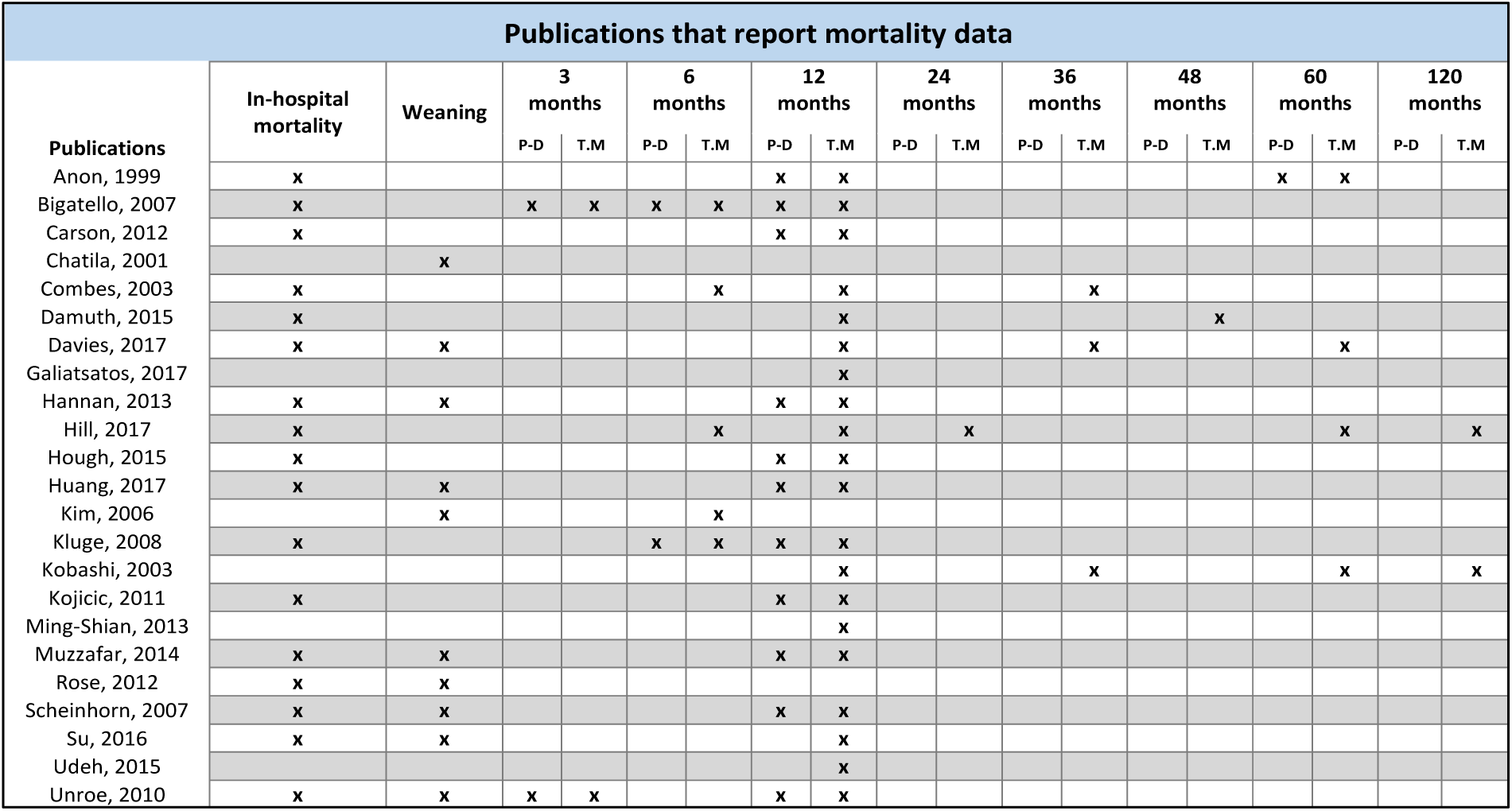
indicates studies that reported mortality outcomes at each timepoint. P.D = post-discharge rates. T.M = total.

Functional data was reported in fifteen papers including discharge/subsequent location, return to work and quality of life ratings. **Table 4a and 4b** highlight results, included patients, and reported publications. The eventual discharge or subsequent location was reported in thirteen publications, with 30.2% of patients being discharged to home either independently or with assistance. Eleven publications reported that 25% of patients were discharged to an acute care facility or skilled nursing home, with eight studies reporting 20.2% of patients discharged to a rehabilitation facility. Functional capacity regarding returning to work was reported in one paper, with 43% of premorbid working patients left disabled and unable to return to work following PMV.

**Table 4a.** highlights functional, psychiatric, and physiological outcomes from included studies.

| Characteristic | Proportion | Reported In:<br>Studies (included patients) |
| --- | --- | --- |
| **Weaning from PMV** |  |  |
| Successful weaning | 57% | 10 (2,970) |
| **Discharge** |  |  |
| Discharged from ICU or equivalent | 73% | 17 (334,524) |
| **Discharge Location** |  |  |
| Home independently or with assistance | 30.2% | 13 (16,831) |
| Acute care facility, skilled nursing home | 25% | 11 (8,473) |
| Rehabilitation facility | 20.2% | 8 (8,293) |
| **Functional capacity** |  |  |
| Pre- morbidly working, left disabled and unable to work | 43% | 1 (25) |
| **Psychiatric** |  |  |
| Post-traumatic stress disorder (PTSD) | 9% | 1 (23) |
| Anxiety | 39% | 1 (23) |
| Depression | 9% | 1 (23) |
| Delirium | 26% | 1 (34) |
| **Ventilator acquired pneumonia (VAP)** |  |  |
| New onset VAP | 25.6% | 3 (313 patients) |

**Table 4b.**
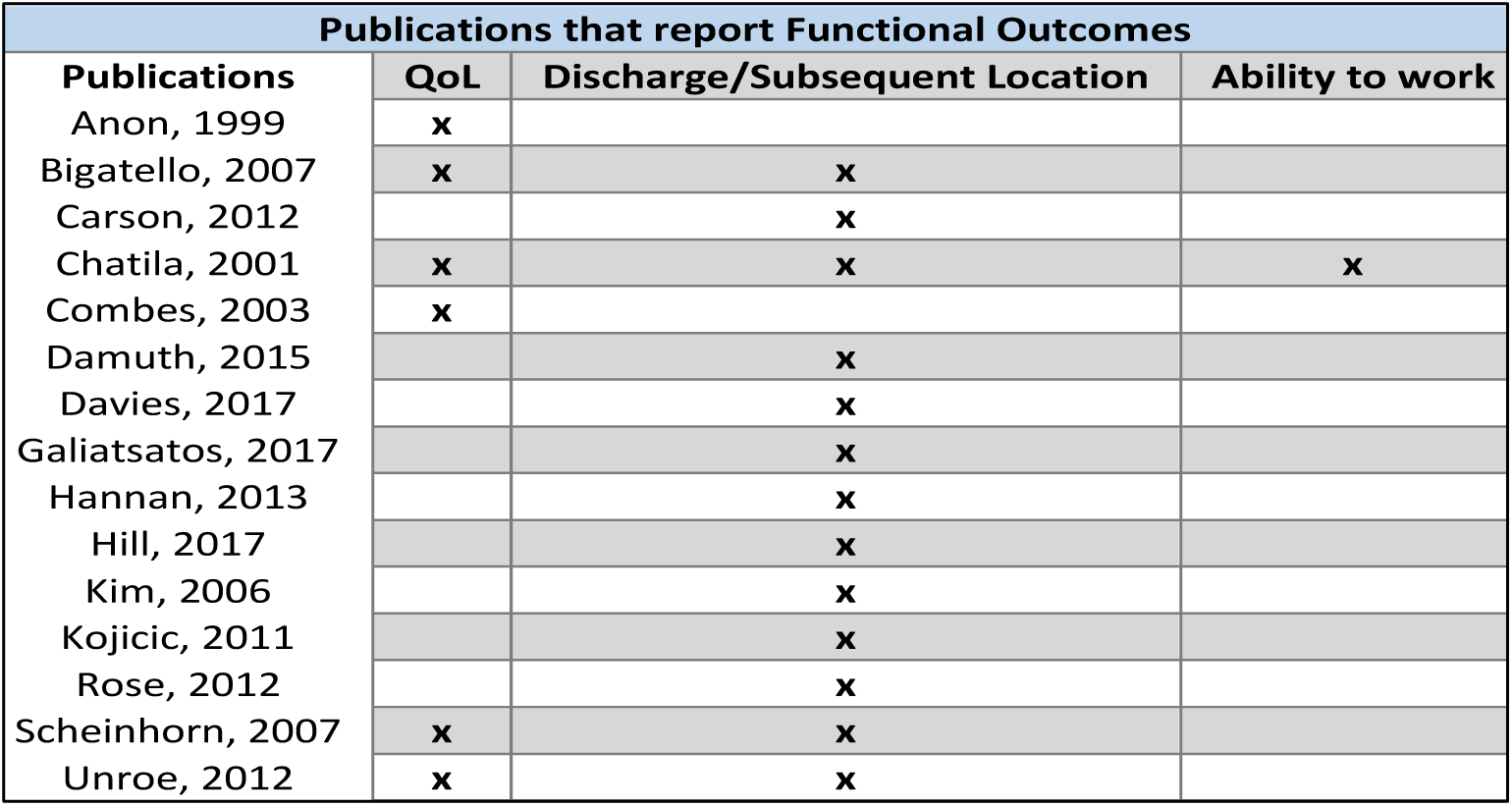
highlights studies that reported functional outcomes. QoL = quality of life.

Quality of life was reported in 6 papers, using varying methods of assessment and at multiple differing timepoints. Objective measures such as ADLs, French Nottingham Health Profile, Zubrod score and Sickness Impact Profile (SIP) along with subjective measures of functional capacity were used. The French Nottingham Health profile completed 3 years after ICU discharge by 89 survivors reported decreased health-related quality of life across physical mobility, emotional reactions, pain, sleep, energy, and global score. The Zubrod score at discharge performed on 831 patients highlighted 69% had poor functional status, while follow up at 12 months of 299 patients demonstrated that 40.1% of patients were still experiencing a poor functional status. A SIP score of ≥ 30 highlights severe function limitation, while a score of ≤5 is seen in healthy adults. 25 patients completed the questionnaire at a median time of 25 months post-ICU discharge, with a mean global SIP of 12 +/- 10, a physical score of 12 +/- 12 and psychosocial score of 9 +/-11. The mean ADL score for a population of 210 patients was 54 +/- 21 and 64 +/- 22 at 3 and 6 months respectively post discharge, indicating a high level of assistance required. Subjective assessment looked at good, fair and poor outcomes, where a good outcome was a patient alive with no functional dependencies, a fair outcome was alive with some functional dependencies and a poor outcome was either alive with complete functional dependencies or dead. In one study with a population of 90 patients, outcomes were assessed at 3 and 12 months, with 37.5% (21% alive and 16.5% dead) having a poor outcome at 12 months, and only 18% of patients improving or sustaining a good outcome from their 3-month assessment. Moreover, in a small population of 5 survivors at 1 year post discharge from PMV, only 2 patients maintained their premorbid good functional level, while the remaining survivors demonstrated an absence of pain, but total disability, with a marked limitation defined as domiciliary confinement except for short, assisted walks.

Psychiatric outcomes were reported in two papers highlighted in Table 4c. PTSD was reported in 9% of patients, assessed using an Impact of events scale-revised score >33 (IES-R). Anxiety, as assessed by the Hospital Anxiety Depression scale (HADS) -anxiety > 11 was seen in 39% of patients, with those patients statistically more likely to have delusional memories of their experience (p=0.008). Depression was reported in 9% of patients as assessed by HADS-depression rating >11.

**Table 4c.** highlights studies that reported psychiatric and physiological outcomes.

| Publications reporting Psychiatric and Physiological Outcomes |  |  |  |  |  |
| --- | --- | --- | --- | --- | --- |
| Publications | PTSD | Anxiety | Depression | Delirium | VAP |
| Huang, 2017 |  |  |  |  | x |
| Kobashi, 2003 |  |  |  |  | x |
| Muzzafar, 2014 |  |  |  | x | x |
| Rose, 2014 | x | x | x |  |  |

Physiological outcomes were reported in three papers outlined in Table 4c. New onset delirium following successful weaning was reported in 26% of patients. While VAP, was reported in 33% of patients.

## DISCUSSION

As IMV continues to become a cornerstone of ICU intervention (Rose and Gerdtz 2009; Mehta et al. 2015; Lone and Walsh 2011; Damuth et al 2015; Zisk-Rony et al. 2019) especially in the age of Covid-19, an understanding of long-term patient outcomes from those that receive PMV has crucial implications on its use in patient management. Moreover, gaining a truer understanding of how these patients recover allows for targeted interventions to reduce ensuing morbidity and the burden placed on families, carers, and the healthcare system more broadly.

IMV of greater than 14 days was used to define PMV for several reasons. Firstly, a recent review of 124 studies (Damuth et al. 2015) used a similar definition. Secondly, clinical consensus of local intensive care specialists at a large Australian tertiary hospital where the authors were based concluded >14 days to be the most appropriate clinical definition. Finally, it was thought that defining PMV as >14 days, rather than any longer period would increase the number of included studies thus improving the accuracy of our findings. Our search was limited to publications within the last 20 years to reduce confounding of results from older studies prior to innovations in critical care practice that have improved survival (Dettmer et al. 2017).

With regard to survival rates, this review highlights that while most patients did survive to hospital ward discharge, a much smaller proportion, less than half, survived beyond 12 months of their discharge, and approximately two thirds were deceased by 48 months. These rates were slightly improved on previously published data which has reported mortality rates as high as 63% (Ming-Shian 2012) at 12 months, highlighting improvements in outcomes with continued utilization of IMV (Mehta et al. 2015). Publication data as well as patient numbers were lacking over some of the other time points, for example only 2 publications reported survival rates at 3 months, and only 5 at 6 months, making it difficult to accurately assess and predict survival rates of patients undergoing PMV.

Functionally, in order for patients to return to their premorbid function, they must not only survive to ICU discharge but be successfully liberated from mechanical ventilation, with only just over half of the patients successfully weaned from PMV, it is not at all surprising that very few patients returned home or to their premorbid state.

Moreover, VAP, a major physiological burden was noted to occur in approximately one quarter of patients. VAP is a well recognised precipitate of chronic critical illness (CCI), a state in which patients are dependent on mechanical ventilation beyond 14 days but are neither improving nor imminently declining, leading to significant morbidity and mortality, including functional dependence, and psychological, cognitive and communication deficits (Meegan et al. 2018; Nelson et al. 2004). With 43% of patients failing to be successfully weaned, this systematic review provides further evidence to the considerable presence of CCI in PMV patients. With this understanding, it is clear why a large proportion of patients required extended, high-acuity care, often in long-term acute facilities, rehabilitation centers and hospices.

Finally, the psychiatric burden of undergoing PMV cannot be overlooked. With our results highlighting up to 39% of patients experiencing psychiatric symptoms and previous studies indicating it could be as high as 74% of patients (Cox et al. 2012), there is a necessity for further investigation, intervention, and follow-up psychiatric care strategies for PMV patients following possible extubation and discharge. Additionally, with QoL scores demonstrating a poor functional status in PMV post 6-12 months post discharge, further research, and consideration of outcomes following PMV must extend beyond merely survival and include the functional burden on life following PMV and hospital discharge.

### Strengths & Limitations

This systematic review provides a broad understanding of the long-term effects of PMV beyond simply survival and demonstrates potentially significant burden to patients and their families. This review has the potential to better inform clinical decision making.

We acknowledge several limitations of our study. Patient-level data was not reported in included papers and therefore analysis could only be performed on mean or pooled data with specific co-morbidities or illness severities not accounted. Additionally, the heterogeneity of the data may lead to variations conclusions, which were minimised by stratifying outcomes, performing pooled analysis, and reporting both in-hospital and total mortality.

Limited data regarding psychiatric outcomes was available, with no objective measures of neurological functioning or cognition were identified. Functionally, with no universal measure of QoL, conclusions on level of morbidity were complicated. Furthermore, QoL reports were often reported at timepoints where large proportions of the patient population had died or may have been experiencing symptoms of the process of dying, potentially confounding results. Objective measures of functional capacity such as independence, fragility, pulmonary function, and exercise tolerance were limited or non-existent in our findings. No findings of CIPM, ARDS, sepsis or multisystem organ failure were reported. Finally, our review synthesized data from multivariable publications, meaning that while some endpoints have a large volume of included patients, others lead to conclusions that are not statistically significant. Finally, there was a delay in publishing of results from the initial completed search which may result in a primacy bias.

### Future Directions

Despite attempting to answer crucial research questions on the psychiatric, functional, and physiological outcomes of critically ill patients requiring PMV, this systematic review highlights that much remains unknown. Future research is needed to truly understand the plight of these patients, specifically, objective measures of functional status, cognition, neurological status and QoL.

Additionally, longer term physiological assessments, from 12-months to 48-months, are indicated as survival rates continue to improve with innovations in critical care practice. These findings will inform the needs of the growing number of PMV patients following their discharge, ensuring appropriate follow-up care, and minimizing morbidity.

## CONCLUSION

Outcomes are poor for critically ill patients requiring PMV in ICU, a quarter of patients will not survive to hospital discharge and fewer than half of the patients will survive beyond 12-months. Additionally, discharged patients suffer from psychiatric conditions, decreased functional ability and poor physiological outcomes, highlighting a need to extend the focus of care beyond merely hospital survival and successful weaning, to reducing long-term morbidity. Finally, as IMV continues to become a mainstay of ICU intervention, especially in the wake of COVID-19, this data and future research has extreme relevance to long-term public health and patient recovery.

## STATEMENTS AND DECLARATIONS

### Ethics and Dissemination

This study did not involve human participants or unpublished secondary data. As such, approval from the Human Research Ethics Committee was not required

### Patient and public involvement

This work analyses existing research studies, and therefore involves no patients or members of the public.

### Consent for publication

Not applicable

### Availability of data and materials

The datasets used and/or analysed during the current study are available from the corresponding author on reasonable request, with all original data available from sources referenced in **Appendix B.**

### Author contributions

All authors (JL, CH) contributed meaningfully to the preparation, consultation, drafting, screening, extracting, and editing of this systematic review. JL (guarantor) conceived the idea and guided all stages of the research. JL (corresponding author) and CH conceptualized the research questions, core research plan details, and data extraction tool, before preparing the search strategy and undertaking this systematic review. All authors approved the final submitted manuscript after several iterations and rounds of editing and agreed to be accountable for all aspects of this protocol.

## Acknowledgments

We would like to thank Dr. Sam Rudham for his assistance and guidance to both authors throughout the entire research processes, as well as assisting with .

## Guarantor

Dr. Jarryd Ludski is the guarantor

## LIST OF ABBREVIATIONS

IMV: Invasive mechanical ventilation
V/Q: ventilation-perfusion
ICU: intensive care units
PMV: Prolonged mechanical ventilation
PRISMA: Preferred Reporting Items for Systematic Reviews
CCU: coronary care unit
ARDS: Acute Respiratory Distress Syndrome
VAP: Ventilator acquired pneumonia
CIPM: critical illness polyneuropathy and myopathy
QoL: Quality of life
ADLs: activities of daily living
PTSD: Post-traumatic stress disorder
APACHE: Acute Physiology and Chronic Health Evaluation
SIP: Sickness Impact Profile
IES-R: Impact of events scale-revised score
HADS: Hospital Anxiety Depression scale
CCI: chronic critical illness

### Appendix A: Search Design

#### Medline

|  |  |  |
| --- | --- | --- |
| 1 | Intensive Care Units/ | 48996 |
| 2 | Critical Care/ | 48465 |
| 3 | intensive care*.mp. | 156157 |
| 4 | critical care*.mp. | 65057 |
| 5 | intensive care unit*.mp. | 130257 |
| 6 | 1 or 2 or 3 or 4 or 5 | 191268 |
| 7 | Respiration, Artificial/ | 45902 |
| 8 | prolonged mechanical vent*.mp. | 1792 |
| 9 | long term mechanical vent*.mp. | 344 |
| 10 | long-term mechanical vent*.mp. | 344 |
| 11 | 7 or 8 or 9 or 10 | 46964 |
| 12 | long term outcome*.mp. | 48300 |
| 13 | long-term outcome*.mp. | 48300 |
| 14 | SURVIVAL/ or SURVIVAL RATE/ | 164715 |
| 15 | 12 or 13 or 14 | 208850 |
| 16 | 6 and 11 and 15 | 668 |
| 17 | limit 16 to yr="1998 -Current" | 552 |

#### CINAHL

|  |
| --- |
| intensive care units or intensive care or intensive care unit or critical care or critical care unit or critical care units |
| **AND** |
| artificial respiration or prolonged mechanical vent* or long-term mechanical vent* or long term mechanical vent* |
| **AND** |
| long term outcome* or long-term outcome* or survival or survival rate |
| Limit: 1998-current |
| **Total - 658** |

### Appendix B: References of included publications

